# Multiplex plasma profiling of synaptic biomarkers in Alzheimer’s disease using NULISA: early alterations, APOE genotype effects, and pTau217 associations

**DOI:** 10.64898/2026.05.21.26353560

**Authors:** C. Martinuzzo, A. Pilotto, C. Tolassi, M. Sauer, A. L. Benedet, A. Rondina, A. Galli, T. Merati, C. Trasciatti, I. Girotto, G. di Molfetta, I. Pola, K. Tan, W. Traichel, S. Caratozzolo, S. Pelucchi, E. Marcello, F. Gardoni, M. Di Luca, H. Zetterberg, N. J Ashton, A. Padovani

## Abstract

**INTRODUCTION:** Synaptic markers are altered in the CSF of Alzheimer’s disease (AD) patients, but their quantification in plasma remains challenging. We evaluated plasma synaptic markers in MCI and mild AD using the nucleic acid–linked immuno-sandwich assay (NULISA) and their correlation with APOE genotype.

**METHODS:** 272 participants (154 CSF-confirmed AD, 118 controls) underwent plasma assessment with the NULISA CNS panel. A subset (*n*=48) also had CSF measurements. Analyses were adjusted for age, sex, comorbidity, and renal function.

**RESULTS:** NULISA revealed plasma alterations in NPTX2, NPTXR, SNAP-25, and VSNL1 in AD, with SNAP-25 and NPTXR already altered at MCI stage. APOE ε4/ε4 carriers showed higher plasma SNAP-25. Plasma SNAP-25 and NPTXR correlated positively with pTau217. No plasma-CSF concordance was observed.

**DISCUSSION:** NULISA identifies plasma synaptic biomarker alterations in early AD, with APOEε4 influencing SNAP-25 levels. Associations with pTau217 suggest a link between synaptic damage and tau phosphorylation. Longitudinal studies are warranted.

## 1. Introduction

Alzheimer’s disease (AD) is characterized by progressive synaptic dysfunction and loss, which occur early in the disease course and are closely associated with cognitive decline [1,2]. Over the past decade, several synaptic proteins have emerged as promising biomarkers in cerebrospinal fluid (CSF), reflecting synaptic dysfunction in vivo. Among these, synaptosomal-associated protein 25 (SNAP-25), neurogranin, and the pentraxin family have shown diagnostic and prognostic utility in AD [3–6].

Despite these advances, the use of CSF-based synaptic biomarkers is limited by the invasiveness of lumbar puncture, hindering their widespread application in clinical practice and large-scale population screening. In contrast, blood-based biomarkers offer a more accessible and scalable alternative. However, detecting brain-derived synaptic proteins in plasma is technically challenging due to their low abundance and the complexity of the peripheral proteome [7]. As a result, standardized and sensitive methods for assessing synaptic markers in blood are still lacking.

Recent advances in ultra-sensitive immunoassays have begun to overcome these barriers. Notably, plasma SNAP-25 levels have been quantified using single-molecule array (Simoa) technology, showing significant elevation in AD and mild cognitive impairment (MCI), and associations with amyloid pathology and cognitive decline [8–11].

The recent development of ultra-sensitive immunoassay technologies has enabled the quantification of neuronal proteins in plasma with high specificity and sensitivity. Among these, the NUcleic acid Linked Immuno-Sandwich Assay (NULISA) represents a novel approach that combines antibody specificity and repeated washing of the immunocomplexes with nucleic acid signal amplification to detect multiple (>100) central nervous system (CNS)–related proteins [12–14]. Whether this technology can reliably quantify synaptic proteins in plasma from AD patients in a disease-relevant manner remains to be fully investigated.

This study utilized NULISA to analyze synaptic biomarkers in a well-characterized cohort, including patients with MCI and mild AD. The primary objectives were to test i) whether this ultra□sensitive technology could detect differences in plasma synaptic biomarker levels across disease stages; ii) whether these markers are associated with clinical characteristics, genetic risk, and established biomarkers of AD pathology.

## 2. Methods

### 2.1 Patients

We retrospectively selected a cohort of 154 patients (98 females; age 72.1 ± 6.9 (range 51.5–84.0); education 9.1 ± 4.3 years (range 5–20); MMSE score 23.3 ± 4.7 (range 9–30) with mild cognitive impairment (AD-MCI, *n*=104) or mild dementia (AD-DEM, *n*=50) who underwent CSF assessment for standard biomarkers at the outpatient Neurodegenerative Clinic, S.C. Neurology of the ASST Spedali Civili of Brescia, Italy. The internal cut-off value for AD diagnosis was set to Aβ42/p-tau181 ratio >11.1, measured in CSF using the fully automated chemiluminescent enzyme immunoassay platform Lumipulse G600 II® (Fujirebio Europe, Gent, Belgium), as previously reported [15], indicating AD biological profile [16].

At the baseline evaluation, all participants were subjected to neurological, general medical examinations, and an extended battery of neuropsychological tests, including Mini-Mental State Examination (MMSE) and Montreal Cognitive Assessment (MoCA), the Neuropsychiatric Inventory (NPI) and Clinical Dementia Rating Scale (CDR), as well as an evaluation of comorbidity using the cumulative illness rating scale (CIRS). Each participant underwent routine blood analyses and brain magnetic resonance imaging to exclude large cortical infarcts and other non-degenerative causes of cognitive deficits.

### 2.2. Control group

For plasma biomarkers comparisons, we included a healthy control (HC) group of 118 volunteers (mean age 68.8 ± 6.2 years, 64% female 136). The groups were matched for sex, while age and education differed significantly between groups and they were used as covariates in all biomarker analyses. This group of neurologically and cognitively normal individuals was recruited from participants’ caregivers, as part of the Life-BIO cohort. The following exclusion criteria were applied: i) diagnosis of any neurological disorder, ii) presence of subjective cognitive complaints, iii) abnormal neurological examination and MoCA Screening, iv) major psychiatric disorder, v) recent inflammatory events. To rule out prodromal AD in controls group a two–cut-off approach for plasma p-tau217 measured using the fully automated chemiluminescent enzyme immunoassay platform Lumipulse was applied. Individuals with p-tau217 <0.22 pg/mL were classified as biomarker-negative, while values between 0.22 and 0.34 pg/mL were considered indeterminate and excluded from binary analyses [17].

The study was approved by the local ethics committee (DMA study (NP 1471) and Neuromultibio Study (NP5285, approved by the Local Brescia ASST Ethic Comitee on the 10.05.2022) and was performed in conformity with the Helsinki Declaration; informed consent was obtained from each study participant or their legally authorised representative.

### 2.3 Plasma collection and analysis

Blood samples were collected from each participant using 7.5 mL tubes containing K2-ethylenediaminetetraacetic acid (K2-EDTA). The tubes were gently inverted 5 to 10 times to mix the blood and then centrifuged at 2500×g for 10 minutes at room temperature (RT). Next, 0.5mL plasma aliquots were pipetted into polypropylene cryotubes and directly stored at ultra-low temperature freezing (ULTF) −80°C. On the day of analysis, the plasma samples were brought to RT (21–23 °C). Following the manufacturer’s instructions, plasma samples were centrifuged at 2000g for 5 minutes. NULISA assays were performed at the Clinical Neurochemistry Laboratory at Sahlgrenska University Hospital (Gothenburg, Sweden). For the NULISAseq CNS Disease Panel, ∼120 neurodegenerative disease-related targets were quantified through immunocomplex formation associated with DNA reporter molecule ligation. DNA reporter molecules were pooled and amplified by PCR, purified and sequenced on Illumina NextSeq 2000. The sequencing data were processed using the NULISAseq algorithm (Alamar Biosciences) as previously reported [14]. Intraplate normalization was performed by dividing the target counts for each sample well by that well’s internal control counts. Interplate normalization was then performed using interplate control (IPC) normalization, wherein counts were divided by target-specific medians of the three IPC wells on that plate. Data were then rescaled, and log2 transformed to obtain NULISA Protein Quantification (NPQ) units for downstream statistical analysis. Samples were included in the analysis only if their NPQ values exceeded the limit of detection (LOD), as provided by Alamar Biosciences. All sample measurements below the LOD were excluded to ensure reliable quantification. From the full NULISA CNS panel, only core synaptic proteins were included, defined as proteins annotated to pre- or post-synaptic localization or core synaptic processes in SynGO and showing brain-elevated or neuron-enriched expression in the Human Protein Atlas (e.g., SNAP-25, NPTX1-2, NPTXR, neurogranin, VGF, BASP1). Only proteins quantifiable above the limit of detection in >90% of samples were included.

### 2.4 CSF collection and analyses

Following standard operating procedures, in a subset of 48 AD patients, CSF samples (6 to 8 mL) were obtained via lumbar puncture at the L3-L4 or L4-L5 interspace in fasting conditions according to the standardised protocol of the outpatient Neurodegenerative clinic. The CSF was collected using 15 mL sterile polypropylene tubes, then centrifuged to avoid gradient effects and sent directly to the hospital laboratory for routine assessments and Lumipulse CSF core AD markers [15]. This subset of samples also underwent the NULISAseq CNS Disease Panel, as described previously. Additionally, 70 CSF samples were analysed for SNAP-25 using the Single molecule array (Simoa) assay Neurobiorepository and Laboratory of advanced biological markers, University of Brescia and ASST Spedali Civili Hospital (Brescia, Italy).

### 2.5 Statistics

Statistical analysis utilized GraphPad Prism version 10 (GraphPad Software, San Diego, California, USA), with p < 0.05 significance threshold. Outliers were identified and removed using the Tukey method. The inter-quartile range was calculated and any value lying beyond 1.5 × IQR from the first or third quartile was excluded. Normality distribution was evaluated using the Shapiro-Wilk test and Q-Q plots. Depending on data distribution, comparisons between diagnostic groups (AD, HC) were performed using Student’s t-test or Welch’s test for normally distributed variables, and Kruskal–Wallis test with Dunn’s post-hoc correction for non-parametric data. To further evaluate relationships between synaptic plasma biomarkers and plasma pTau217, linear regression analyses were conducted, adjust for potential confounders. Finally, a hierarchical multiple linear regression with backward elimination was performed to identify independent predictors of plasma pTau217. Synaptic biomarkers and relevant covariates were entered sequentially across three models. Model fit was evaluated using ΔR², AIC, and standardized β coefficients.

## 3. Results

### 3.1 Plasma synaptic biomarker levels across disease stages

Comparative analysis of plasma synaptic biomarkers between AD patients and controls revealed distinct group differences across disease stages, independently from age and sex **(Table 1, Supplementary Table 1 and Fig.1).** From the full NULISA CNS panel, only core synaptic proteins were included, defined as proteins annotated to pre- or post-synaptic localization or core synaptic processes in SynGO and showing brain-elevated or neuron-enriched expression in the Human Protein Atlas (**Supplementary Fig. 1s**). Of these core synaptic proteins, only NPTX2, NPTXR, SNAP-25, and VSNL1 showed statistically significant differences between AD and HC (p < 0.05) (**Fig. 1, panel A**). Notably, only SNAP-25 and NPTXR exhibited consistent differences across disease sub-stages (**Fig. 1, panel B**).

**Fig. 1.**
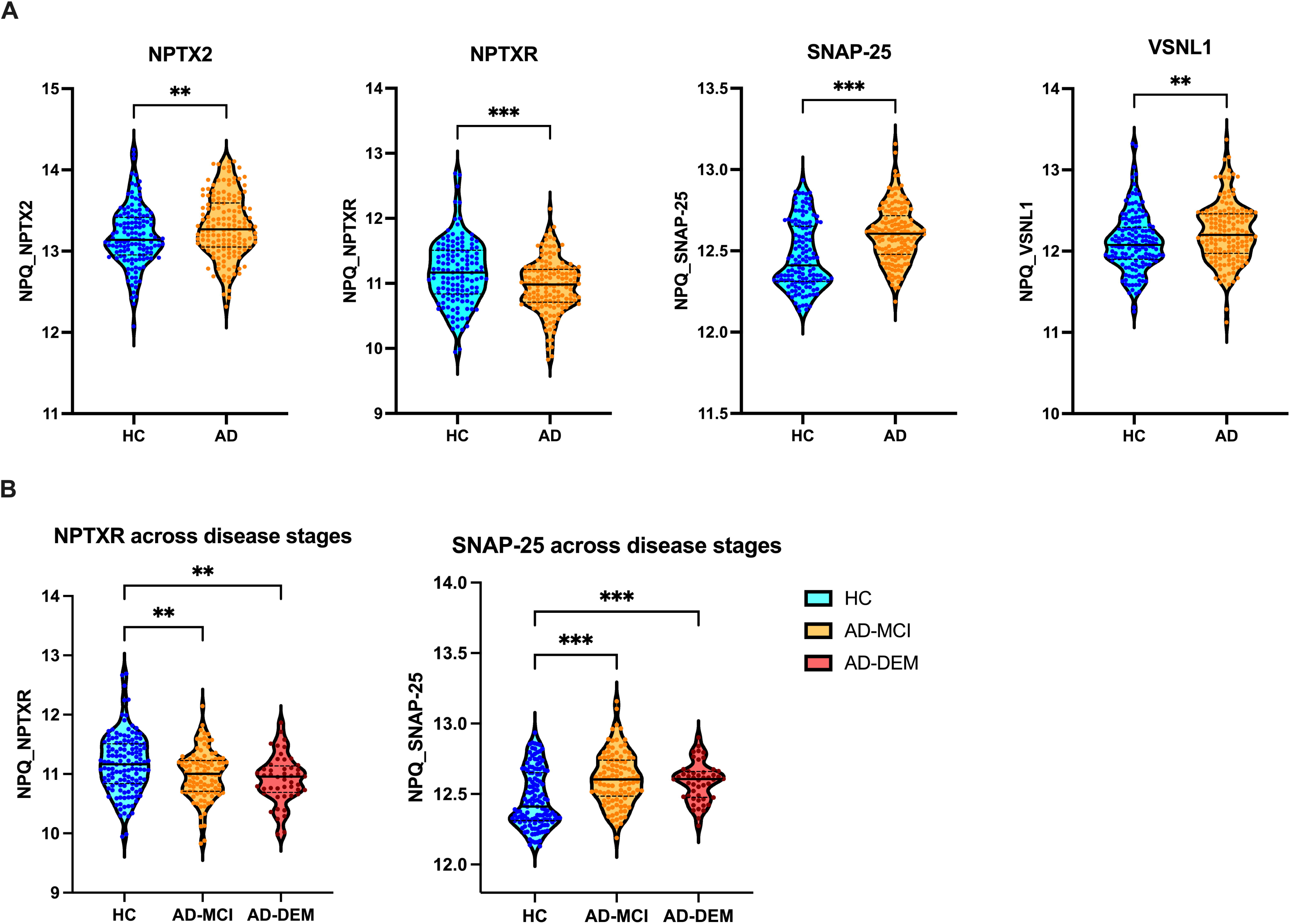
Comparisons of plasma synaptic biomarkers values in Alzheimer’s disease and across disease stages. **(A)** Comparison of NPQ levels of NPTX2, NPTXR, SNAP-25, and VSNL1 between healthy controls (HC) and patients with Alzheimer’s disease (AD) (blue, and orange violin plots respectively). **(B)** NPTXR and SNAP-25 NPQ levels across disease stages: healthy controls (HC), Alzheimer’s disease with mild cognitive impairment (AD-MCI), and Alzheimer’s disease with dementia (AD-DEM) (blue, orange and red violin plots respectively). The violin plots illustrate the median concentration and the 25th and 75 percentiles. Any statistically significant p-values are displayed as following: * p<0.05, ** p<0.01, *** p<0.001. *Abbreviations: NPTX1, Neuronal Pentraxin 1; NPTX2, Neuronal Pentraxin 2; NPTXR, Neuronal Pentraxin Receptor; NRGN, Neurogranin; SNAP-25, Synaptosomal-Associated Protein 25; VSNL1, Visinin-like protein 1; NPQ, NULISA Protein Quantification*.

**Table 1.**
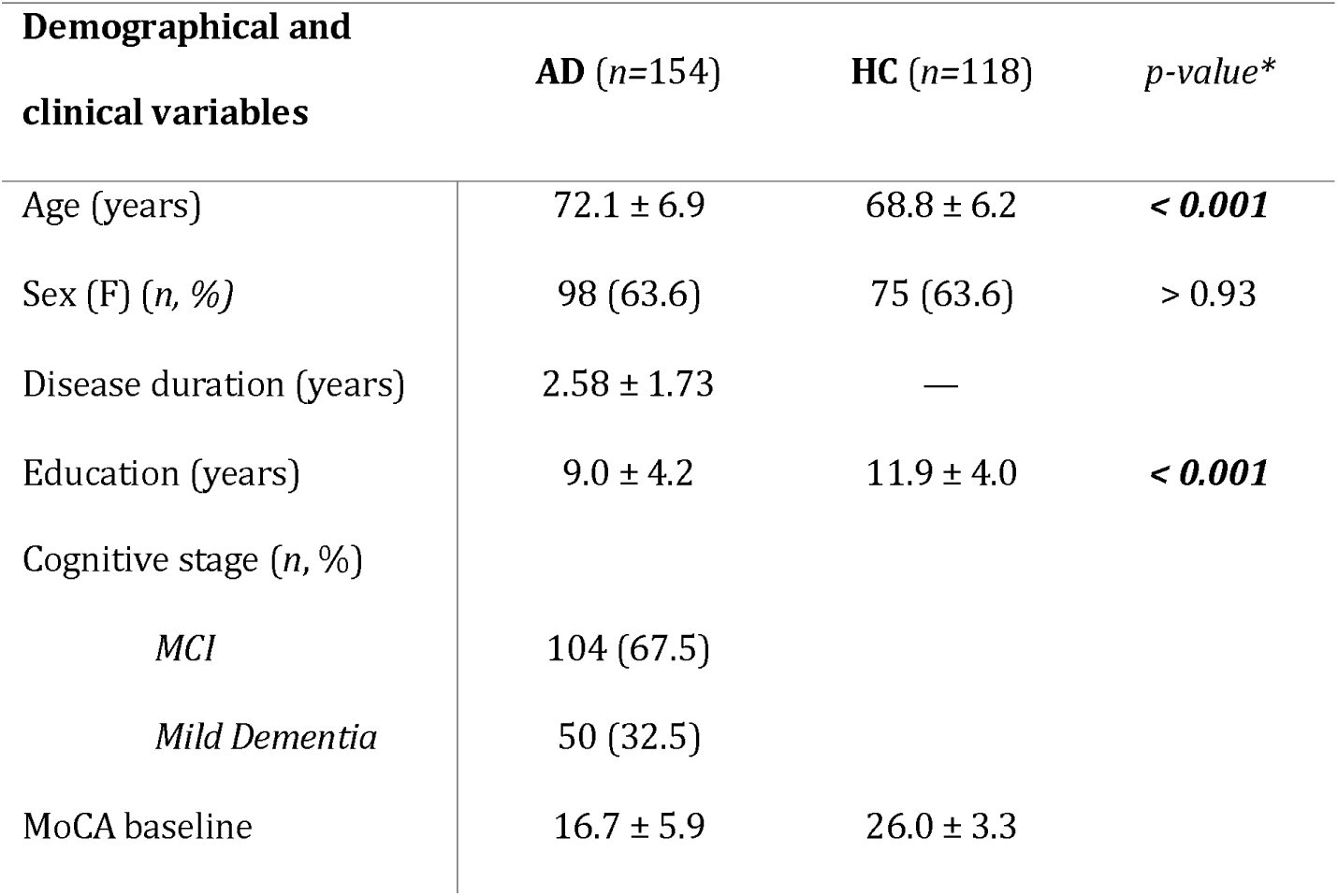
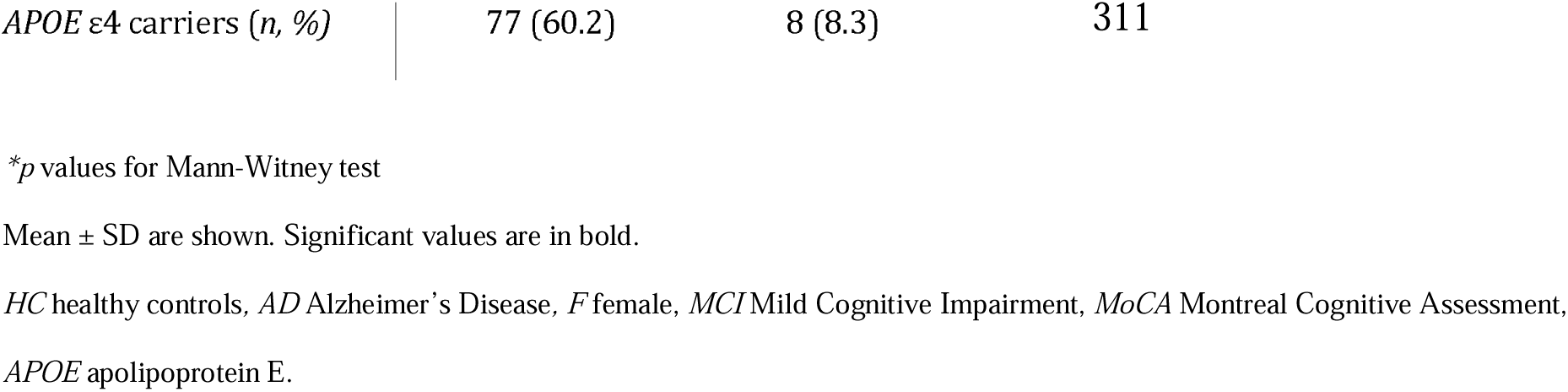
Main clinical characteristics and biomarkers levels of cohort participants.

SNAP-25 levels were significantly higher in AD patients compared with HC (*p* < 0.001), with this difference consistently observed across both the MCI and dementia stages. Conversely, NPTXR levels were significantly reduced in AD relative to controls, with similar levels between MCI and mild dementia groups (*p* < 0.01). None of the other synaptic biomarkers included show significant differences when AD patients were stratified by clinical stages. Within the AD cohort, SNAP-25 and NPTXR levels were not associated with cognitive status, neuropsychiatric symptoms, or short-term disease progression, and were independent of comorbidity burden assessed by the CIRS scale.

### 3.2 Correlations between CSF vs. plasma synaptic biomarkers

In a subset of 48 AD, NULISA was performed in CSF to test CSF/plasma correlations. The distribution of synaptic biomarkers revealed distinct concentration ranges across plasma and CSF, with SNAP-25, NPTX2, NPTXR, and VSNL1 each forming separate clusters. Among these, NPTXR showed the greatest discordance between CSF and plasma values, with minimal overlap, whereas the other biomarkers displayed partially overlapping yet still clearly differentiated distributions between the two matrices **(Supplementary Fig. 2s).** In AD, positive correlations among neuropentraxin family proteins were detected, detailed in supplementary material (supplementary material). To assess assay concordance, we compared plasma and CSF SNAP-25 concentrations measured with NULISA and Simoa. No significant relationship was observed between plasma and CSF SNAP-25 using the same platform (NULISA; R² = 0.026, p = 0.28) (**Supplementary Fig. 5s** - **Panel A**). Likewise, plasma SNAP-25 measured with NULISA did not correlate with CSF SNAP-25 quantified by Simoa (R² = 0.038, p = 0.11) (**Supplementary Fig. 5s** - **Panel B**). In contrast, CSF SNAP-25 concentrations showed a significant correlation using different platforms (R² = 0.23, p < 0.001) (**Supplementary Fig. 5s** - **Panel C**).

### 3.3 Effects of *APOE* on plasma SNAP-25 levels

To investigate whether *APOE* genotype influences plasma synaptic biomarkers levels, we performed a Kruskal–Wallis test followed by Dunn’s multiple comparisons test. A significant overall effect of *APOE* genotype on plasma SNAP-25 was observed. Post-hoc analyses revealed that individuals carrying the ε4/ε4 genotype showed significantly higher SNAP-25 levels compared to both ε3/ε3 (adjusted *p* = 0.0078) and ε3/ε4 carriers (adjusted *p* = 0.0068). No significant difference was detected between ε3/ε3 and ε3/ε4 genotypes (p > 0.05) **(Fig. 2).** In contrast, *APOE* genotype did not have a significant effect on the levels of any of the other synaptic biomarkers analysed (p > 0.05 for all comparisons).

**Fig. 2.**
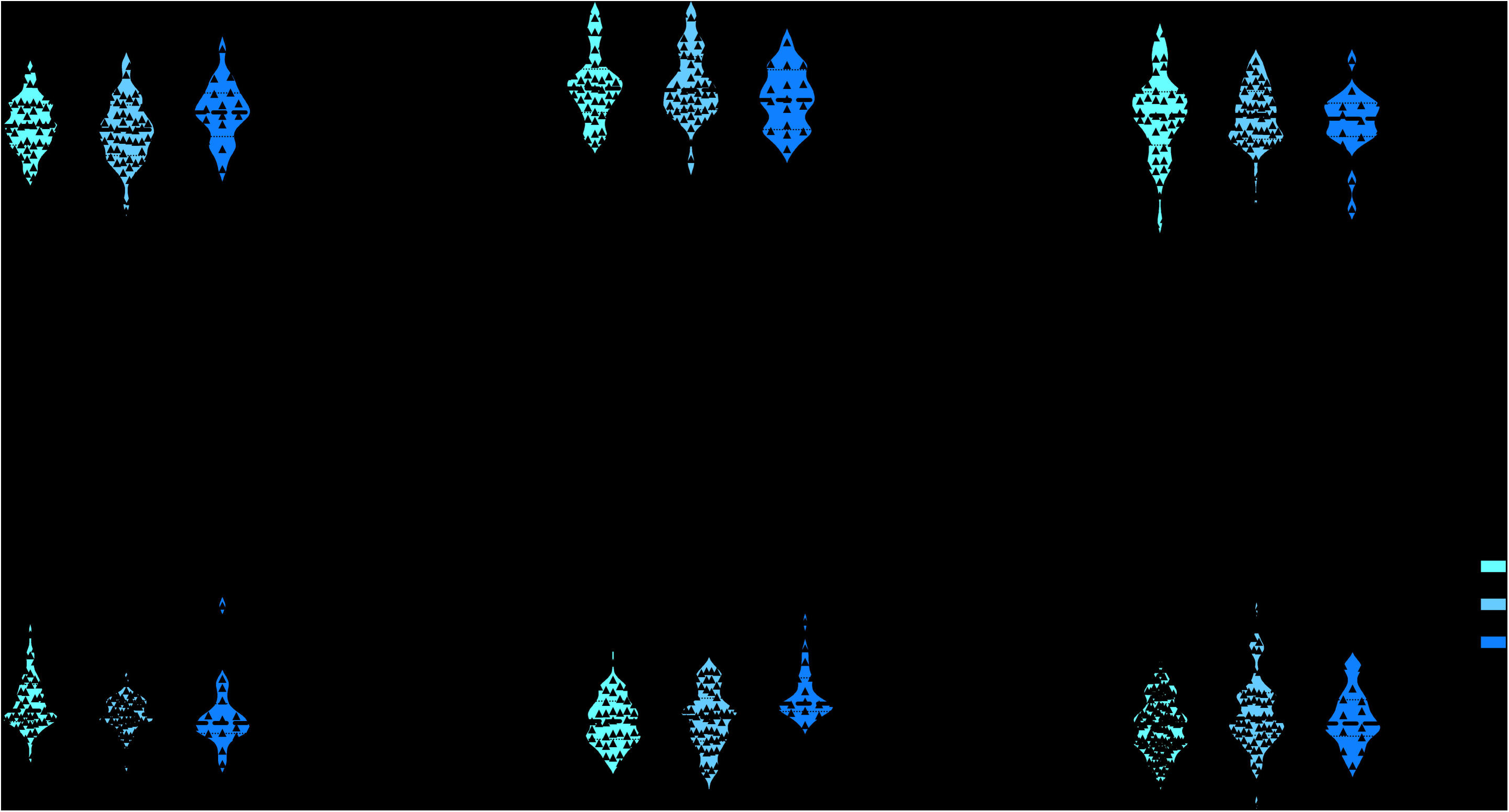
Comparisons of plasma synaptic biomarkers values (namely, NPTX1, NPTX2, NPTXR, NRGN, and SNAP-25) (in NPQ) in ε3/ ε3, ε3/ ε4, and ε4/ ε4 *APOE* genotypes subgroups of AD patients. The violin plots illustrate the median concentration and the 25th and 75 percentiles. Any statistically significant p-values are displayed as following: * p<0.05, ** p<0.01, *** p<0.001. *Abbreviations: NPTX1, Neuronal Pentraxin 1; NPTX2, Neuronal Pentraxin 2; NPTXR, Neuronal Pentraxin Receptor; NRGN, Neurogranin; SNAP-25, Synaptosomal-Associated Protein 25; VSNL1, Visinin-like protein 1; NPQ, NULISA Protein Quantification; APOE, Apolipoprotein E;* ε*3/*ε*3, APOE* ε*3 homozygous genotype;* ε*3/*ε*4, APOE* ε*3/*ε*4 heterozygous genotype;* ε*4/*ε*4, APOE* ε*4 homozygous genotype; AD, Alzheimer’s disease*.

### 3.4 Correlation between SNAP-25, other synaptic biomarkers and pTau217 in plasma

In plasma, SNAP-25 levels showed a significant correlation with NPTX1 (r = 0.33, p<0.001), with no relationship with other synaptic-associated proteins (namely, NPTX2, NPTXR, neurogranin). Moreover, within the neuronal pentraxin family, NPTX1, NPTX2, and NPTXR were weakly intercorrelated (r = 0.158–0.242, p < 0.01; supplementary material for additional correlations). As a core plasma biomarker of AD pathology, NULISA p-tau217 was selected as the dependent variable to assess its relationship with synaptic proteins. Among the proteins analysed, only SNAP-25 and NPTXR showed significant positive correlations with pTau217, although the proportion of explained variance was modest (SNAP-25: R² = 0.085, p = 0.0003; NPTXR: R² = 0.058, p = 0.0027). When stratifying patients by disease stage, the positive association between synaptic biomarkers and pTau217 remained significant exclusively in the MCI subgroup, but not in individuals with established dementia **(Fig. 3).** To further explore these associations, a backward stepwise multivariate linear regression was performed including NPTX1, NPTX2, NPTXR, and SNAP-25 as covariates. Model fit was evaluated using AIC and adjusted R². The final model retained only SNAP-25 and NPTXR as independent predictors of p-tau217 (AIC = 254.731; adjusted R² = 0.109; both predictors p < 0.01).

**Fig. 3.**
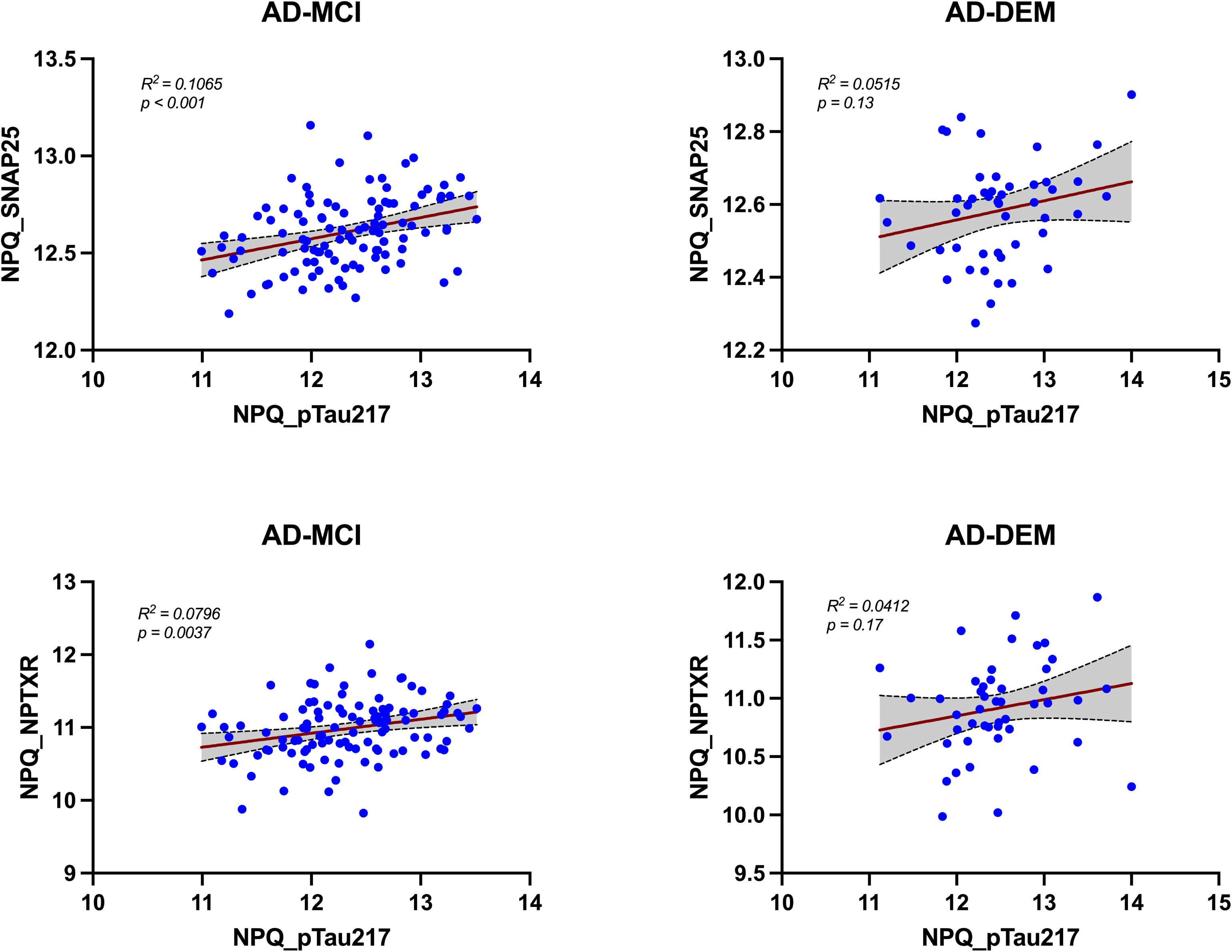
Correlations between plasma pTau217 and plasma synaptic markers. In AD-MCI patients (left panels), pTau217 showed a significant positive correlation with SNAP-25 (R² = 0.1065; p < 0.001) and NPTXR (R² = 0.0796; p = 0.0037). In contrast, correlations in AD-DEM patients (right panels) were weaker and did not reach statistical significance (SNAP25: R² = 0.0515; p = 0.13; NPTXR: R² = 0.0412; p = 0.17). Red lines indicate linear regression fits with 95% confidence intervals (grey shading). *Abbreviations: pTau217, tau phosphorylated at threonine 217; SNAP25, Synaptosomal-Associated Protein 25; NPTXR, Neuronal Pentraxin Receptor; AD-MCI, Alzheimer’s disease at the mild cognitive impairment stage; AD-DEM, Alzheimer’s disease dementia stage*.

## 4. Discussion

The present study demonstrated significant alterations in plasma synaptic biomarkers across the AD continuum using the new NULISA platform, highlighting the potential of SNAP-25 and NPTXR as a translational biomarker detectable in plasma.

Consistent with prior studies, we observed significantly elevated levels of SNAP-25 in plasma among AD subjects, supporting its role as a presynaptic dysfunction marker that can be captured in peripheral fluids and reflects early synaptic degeneration in AD pathology [6,18–20].

The stratification by *APOE* genotype revealed that homozygous ε4/ε4 carriers exhibit significantly higher plasma SNAP-25 than ε3/ε3 or even ε3/ε4 carriers. This finding aligns with the well-documented role of the *APOE* ε4 allele as a major genetic risk factor for AD and its influence on synaptic pathology [21, 22], and with more recent finding showing that plasma SNAP-25 levels were significantly higher levels in *APOE* ε4 vs ε3 carriers [23]. The elevated plasma SNAP-25 in ε4 homozygotes may reflect increased synaptic degeneration or altered synaptic protein turnover associated with the ε4 allele’s pathogenic mechanisms. Interestingly, no significant effects of *APOE* genotype were detected on other synaptic biomarkers, including NPTX1, NPTX2, NPTXR, or neurogranin. This specificity suggests that SNAP-25 may be particularly sensitive to *APOE*-related synaptic changes, supporting a gene-dose effect of *APOE* ε4 in exacerbating synaptic vulnerability, consistent with prior CSF and imaging studies linking *APOE* ε4 to accelerated synaptic loss and amyloid burden [24,25].

In addition, plasma VSNL-1 levels were significantly higher in AD compared to controls, supporting its role as a marker of neuronal injury detectable in peripheral blood. This finding is consistent with prior evidence showing increased VSNL-1 in CSF in AD [26,27] and with more recent studies demonstrating elevated blood levels using ultrasensitive assays [28], suggesting that VSNL-1 primarily reflects neurodegeneration rather than disease-specific pathology.

NPTXR showed an opposite trend, with lower levels in AD patients compared to the HC group. These results are in line with recent findings showing decreased CSF NPTXR correlates with neurodegeneration and clinical progression in AD [29–31]. Moreover, the dynamics of NPTX2 levels in the AD group compared to controls reached statistical significance only when AD patients were considered as a single group, potentially due to low statistical power. An interesting observation is their relatively higher value in dementia stages, at variance with data in CSF [32–34].

Given these compartment-specific differences, examining the CSF associations among synaptic proteins becomes essential to contextualize the plasma findings. In the CSF compartment, the strong correlations observed among NPTX1, NPTX2, and NPTXR highlight their shared role in synaptic function and suggest that alterations in this protein family may represent a convergent signature of synaptic integrity [35]. Their associations with SNAP-25 further support the concept that multiple synaptic compartments are co-affected in disease, pointing to a coordinated vulnerability of pre- and postsynaptic elements [36].

The absence of significant correlations between plasma and CSF synaptic markers illustrates the challenge of translating central nervous system neurodegenerative signals into peripheral blood measurements, likely reflecting factors such as blood–brain barrier permeability and peripheral metabolism. Consistent with this interpretation, previous findings reported that CSF and plasma SNAP-25 showed only a weak positive correlation, while VAMP2 levels in the two compartments were not related at all [8]. These findings highlight an important clinical implication: while blood-based synaptic biomarkers offer practical advantages, their interpretation must be cautious and ideally supported by CSF data. In line with this, our cross-platform comparison further demonstrates that concordance emerges only when measurements are performed within the same biological matrix, whereas plasma SNAP-25 does not merely reflect CSF concentrations regardless of the analytical platform employed, consistent with recent reports showing that agreement between NULISA and Simoa is generally robust only when assays are applied to the same sample type or biological compartment [12, 40].

In plasma, the selective correlation between SNAP-25 and NPTX1 is of particular interest, in line with their involvement in presynaptic release mechanisms and stabilization of trans-synaptic complexes. In line with this, no correlations with SNAP-25 were observed for neurogranin or NPTXR, which reflects postsynaptic dendritic signalling rather than presynaptic integrity [35,37]. Notably, we also identified a modest but significant associations of both plasma SNAP-25 and NPTXR with plasma pTau217, the best AD pathology diagnostic marker which has also an important prognostic value at individual level [38]. This might link synaptic dysfunction directly with abnormal tau phosphorylation, even indirectly from the neurodegenerative process [39]. Interestingly, the associations between synaptic biomarkers and pTau217 were significant in the MCI subgroup but not in established dementia. This might reflect temporal dynamics: synaptic dysfunction and (mis)compensatory mechanisms may be more pronounced, or more detectable, in the earlier disease stages. This pattern aligns with models suggesting synaptic biomarkers may rise early in response to emerging tau phosphorylation even before the whole neurodegenerative process occurs [36,40], thus representing a promising biological hallmark of the individual brain resilience. A further possibility is that synaptic and tau biomarkers increase in parallel during the early phases of AD, reflecting concomitant, though not necessarily causally linked, processes within the evolving pathophysiological cascade; ongoing longitudinal studies will be crucial to further clarify the temporal and mechanistic relationships between these markers.

The detection of SNAP-25 and NPTXR alterations in plasma of AD using the automated NULISA platform open new scenarios in individual monitoring and biological subtyping of AD spectrum. The modulation of SNAP-25 by *APOE* further suggests that personalized biomarker profiling could enhance disease monitoring and therapeutic stratification. While the lack of correlation between plasma and CSF biomarkers reflects methodological and physiological barriers, the observed associations with plasma pTau217 further support the relevance of peripheral synaptic markers in capturing early synaptic (mal)adaptive mechanisms both in presynaptic and postsynaptic compartments. Future longitudinal studies should investigate the dynamic changes of these biomarkers in relation to cognitive decline, imaging findings, and response to emerging disease-modifying treatments.

## Data Availability Statement

The data supporting the findings of this study are available from the corresponding author upon reasonable request, subject to institutional and ethical restrictions.

## Acknowledgements

The study has been supported by the European Union - Next Generation EU - NRRP M6C2 - Investment 2.1 Enhancement and strengthening of biomedical research in the NHS– PNRR-Health PNRR-MAD-2022-12376110.

## Author contributions

AP and APi contributed to the conceptualization and design of the study. AP, CM and APi and contributed to drafting the text or preparing the figures. CM, CTo, TM, IG, SC contributed to data acquisition. MS, ALB, AR, AG, CT, GdM, IP, KT, WT, SP, EM, FG, MdD, HZ, NJA commented and revised the manuscript.

