## Supplementary Materials for "Multiplex plasma profiling of synaptic biomarkers in Alzheimer’s disease using NULISA: early alterations, APOE genotype effects, and pTau217 associations"

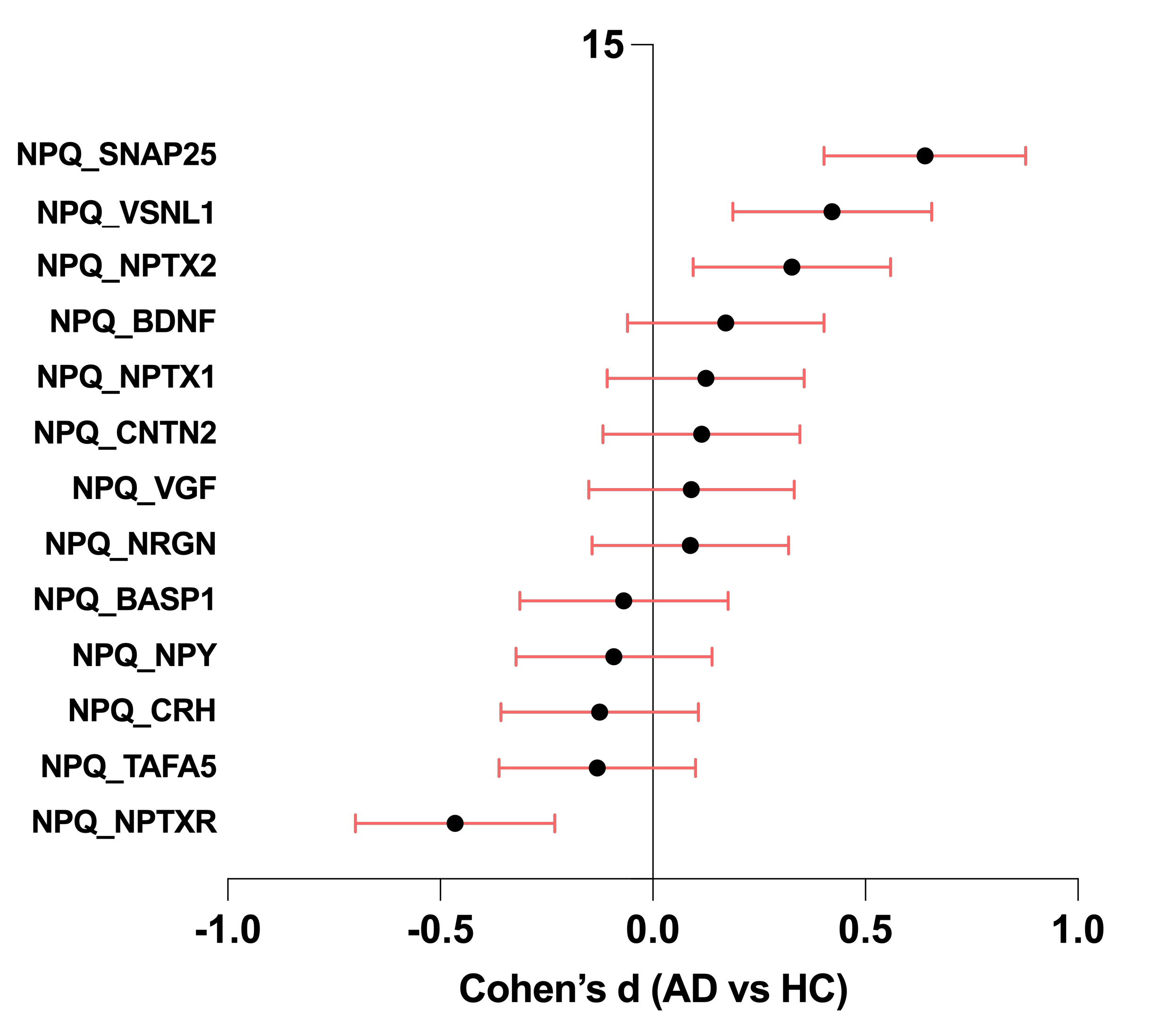

**Supplementary Fig. 1s.** Forest plot showing standardized mean differences (Cohen’s d) between Alzheimer’s disease (AD) patients and healthy controls (HC) for synaptic proteins included from the NULISA CNS panel. Points represent effect sizes and horizontal bars indicate 95% confidence intervals. Positive values indicate higher levels in AD, while negative values indicate lower levels compared to controls. A vertical line at zero denotes no difference between groups.

*Abbreviations: BASP1 Brain Abundant Membrane Attached Signal Protein 1, BDNF Brain-Derived Neurotrophic Factor, CNTN2 Contactin-2, CRH Corticotropin-Releasing Hormone, NRGN Neurogranin, NPTX1* neuronal pentraxin 1, *NPTX2* neuronal pentraxin 2, *NPTXR* neuronal pentraxin receptor*, NPY Neuropeptide Y, SNAP-25* synaptosomal-associated protein 25, *TAFA5 TAFA Chemokine-Like Family Member 5, VGF Nerve Growth Factor Inducible, VGF Nerve Growth Factor Inducible, VSNL1 Visinin-Like Protein 1; NPQ, NULISA Protein Quantification*

**Supplementary Table 1.**

| NULISA plasma synaptic biomarkers | AD (*n=*154) | HC (*n=*118) | *p* value* | *Post-hoc comparisons^†^* |
| --- | --- | --- | --- | --- |
| BASP1 (NPQ) | 12.5 ± 1.6 | 12.6 ± 1.7 | 0.814 | *AD-DEM < HC p>0.05*  *AD-MCI < HC p>0.05* |
| BDNF (NPQ) | 14.0 ± 1.9 | 13.9 ± 1.7 | 0.250 | *AD-DEM < HC p>0.05*  *AD-MCI < HC p>0.05* |
| CNTN2 (NPQ) | 8.1 ± 1.4 | 7.9 ± 1.7 | 0.248 | *AD-DEM < HC p>0.05*  *AD-MCI < HC p>0.05* |
| CRH (NPQ) | 12.6 ± 1.1 | 12.7 ± 1.0 | 0.092 | *AD-DEM < HC p>0.05*  *AD-MCI < HC p>0.05* |
| NRGN (NPQ) | 14.2 ± 1.9 | 14.0 ± 1.7 | 0.133 | *AD-DEM < HC p>0.05*  *AD-MCI < HC p>0.05* |
| NPTX1 (NPQ) | 11.8 ± 0.5 | 11.7 ± 0.5 | 0.657 | *AD-DEM < HC p>0.05*  *AD-MCI < HC p>0.05* |
| NPTX2 (NPQ) | 13.3 ± 0.4 | 13.2 ± 0.4 | ***0.027*** | ***AD-DEM < HC p<0.05***  ***AD-MCI < HC p<0.05*** |
| NPTXR (NPQ) | 11.0 ± 0.4 | 11.2 ± 0.5 | ***< 0.001*** | ***AD-DEM < HC p<0.001***  ***AD-MCI < HC p<0.001*** |
| NPY (NPQ) | 15.0 ± 1.8 | 15.1 ± 1.5 | 0.857 | *AD-DEM < HC p>0.05*  *AD-MCI < HC p>0.05* |
| SNAP-25 (NPQ) | 12.6 ± 0.2 | 12.5 ± 0.2 | ***< 0.001*** | ***AD-DEM < HC p<0.001***  ***AD-MCI < HC p<0.001*** |
| TAFA5 (NPQ) | 10.9 ± 1.1 | 11.0 ± 0.6 | 0.528 | *AD-DEM < HC p>0.05*  *AD-MCI < HC p>0.05* |
| VGF (NPQ) | 12.7 ±1.7 | 12.6 ± 2.0 | 0.249 | *AD-DEM < HC p>0.05*  *AD-MCI < HC p>0.05* |
| VSNL1 (NPQ) | 12.2 ± 0.4 | 12.1 ± 0.3 | ***< 0.001*** | *AD-DEM < HC p>0.05*  ***AD-MCI < HC p<0.05*** |

*Abbreviations: BASP1 Brain Abundant Membrane Attached Signal Protein 1, BDNF Brain-Derived Neurotrophic Factor, CNTN2 Contactin-2, CRH Corticotropin-Releasing Hormone, NRGN Neurogranin, NPTX1* neuronal pentraxin 1, *NPTX2* neuronal pentraxin 2, *NPTXR* neuronal pentraxin receptor*, NPY Neuropeptide Y, SNAP-25* synaptosomal-associated protein 25, *TAFA5 TAFA Chemokine-Like Family Member 5, VGF Nerve Growth Factor Inducible, VGF Nerve Growth Factor Inducible, VSNL1 Visinin-Like Protein 1; NPQ, NULISA Protein Quantification*

^*^ *p* values Kruskal–Wallis test

*^†^* *p* values for Dunn’s post-hoc test

Mean ± SD are shown.

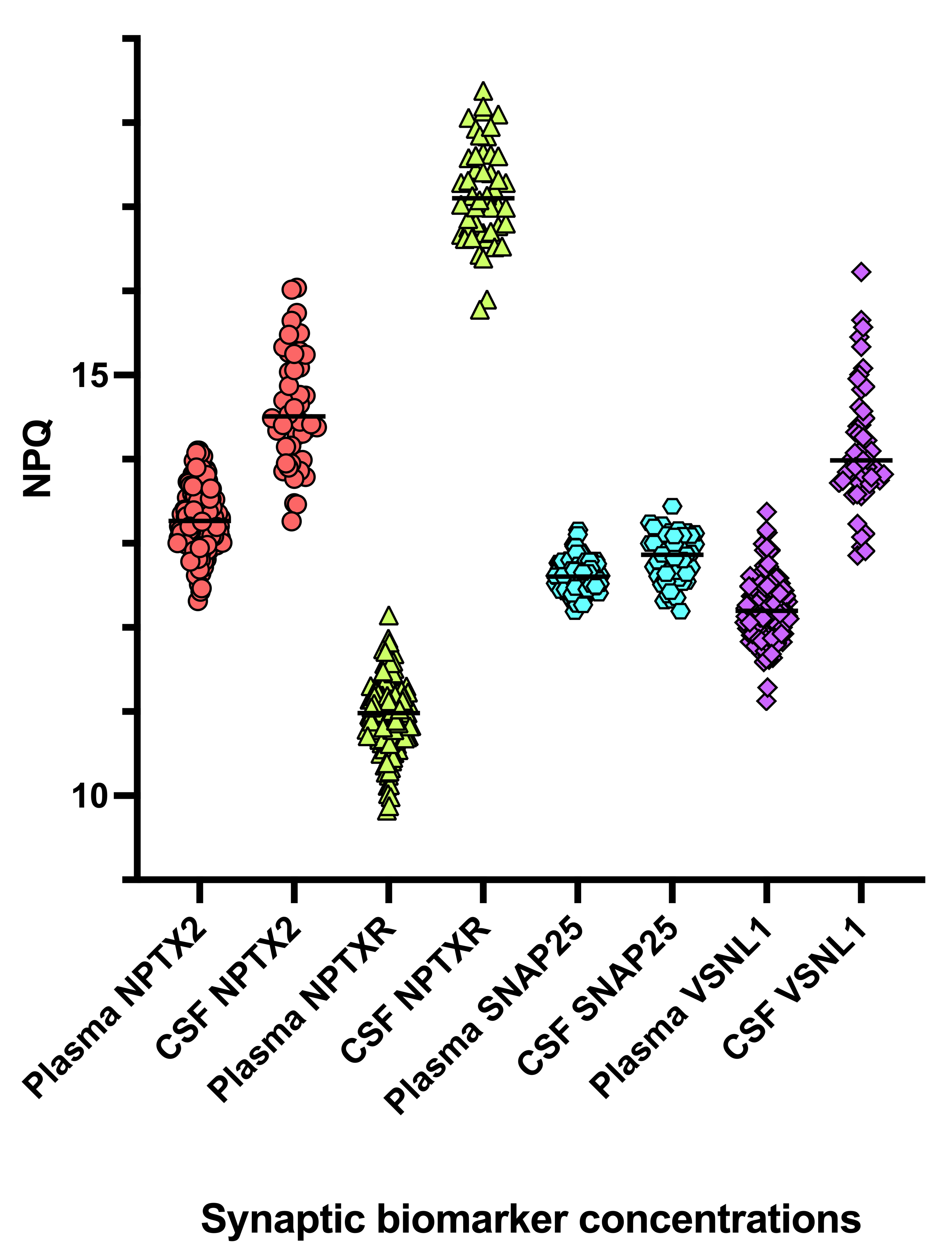

**Supplementary Fig. 2s.** Distribution of core synaptic biomarker concentrations in plasma and CSF.

Dot plots showing the distribution of NPTX2, NPTXR, SNAP-25, and VSNL1 measured in plasma and CSF. Each biomarker displays a distinct concentration range across the two matrices.

*Abbreviations: NPTX1, Neuronal Pentraxin 1; NPTX2, Neuronal Pentraxin 2; NPTXR, Neuronal Pentraxin Receptor; NRGN, Neurogranin; SNAP-25, Synaptosomal-Associated Protein 25; VSNL1, Visinin-like protein 1; NPQ, NULISA Protein Quantification*

**Correlations between synaptic proteins**

In plasma, within synaptic biomarkers, NPTXR was significantly associated with NPTX2 (r=0.24, p=0.003), SNAP25 (r=0.18, p=0.026), and VSNL1 (r=0.21, p=0.009) **(Supplementary Fig. 3s, panel A)**; it also demonstrated significant positive correlations with NfL, GFAP, and pTau217 (r≈0.22–0.28, all p≤0.005), but not with Aβ42/Aβ40. SNAP-25 levels showed a significant correlation with NPTX1 (r = 0.33, p<0.001), with no relationship with other synaptic-associated proteins (namely, NPTX2, NPTXR, NRGN) **(Supplementary Fig. 4s).**

In CSF, NPTXR showed strong correlations within the synaptic panel, being highly associated with NPTX2, SNAP25, and VSNL1 (r=0.68–0.93, all p<0.001), and further correlated with NfL and GFAP (r≈0.48–0.61, p<0.001), but not with pTau217 or Aβ42/Aβ40. Similarly, SNAP25 was strongly associated with the other synaptic markers (r≈0.61–0.74, all p<0.001) and showed robust correlations with NfL and GFAP (r=0.60–0.81, p<0.001), while it was not associated with pTau217 but displayed a modest correlation with Aβ42/Aβ40 (r=0.42, p=0.003) **(Supplementary Fig. 3s, panel B).**

Finally, no significant correlations were observed between the CSF synaptic biomarkers and their plasma counterparts **(Supplementary Fig. 3s, panel C).**

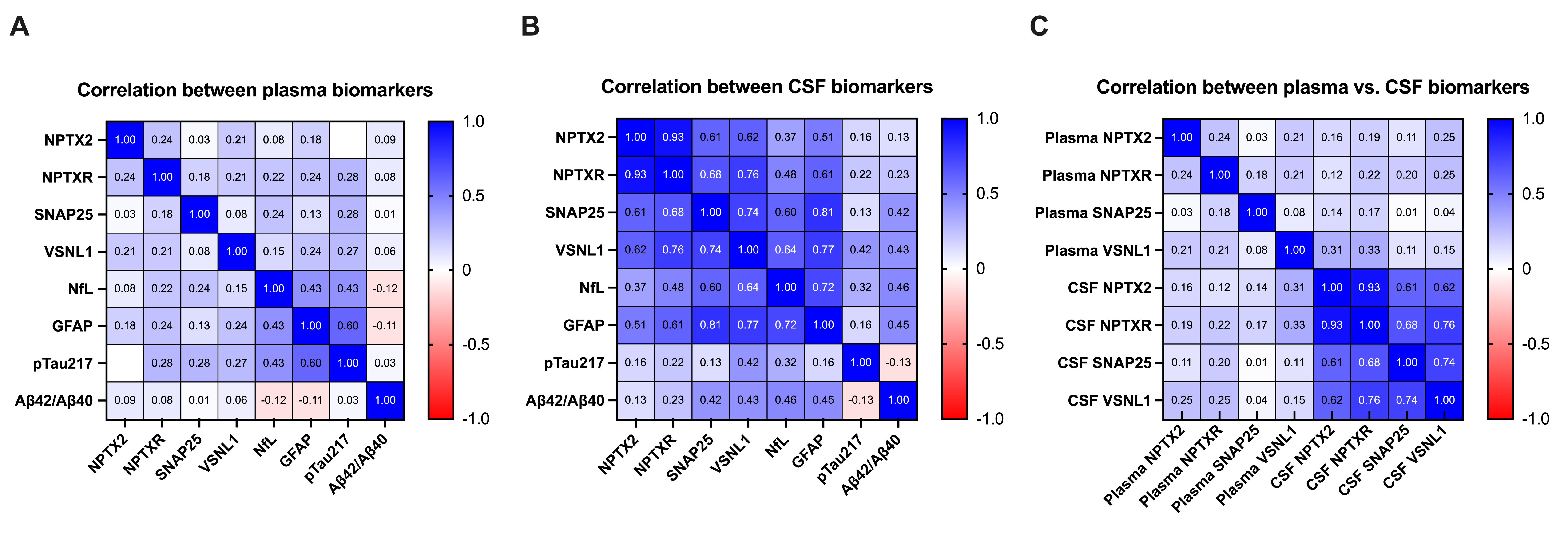

**Supplementary Fig. 3s.** Correlation matrix of core synaptic and neurodegeneration-related biomarkers measured in cerebrospinal fluid (CSF) and plasma. The heatmap displays Sperman correlation coefficients ranging from –1.0 (strong negative correlation, red) to +1.0 (strong positive correlation, blue).

*Abbreviations: NPTX1, Neuronal Pentraxin 1; NPTX2, Neuronal Pentraxin 2; NPTXR, Neuronal Pentraxin Receptor; NRGN, Neurogranin; SNAP-25, Synaptosomal-Associated Protein 25; VSNL1, Visinin-like protein 1; NPQ, NULISA Protein Quantification*

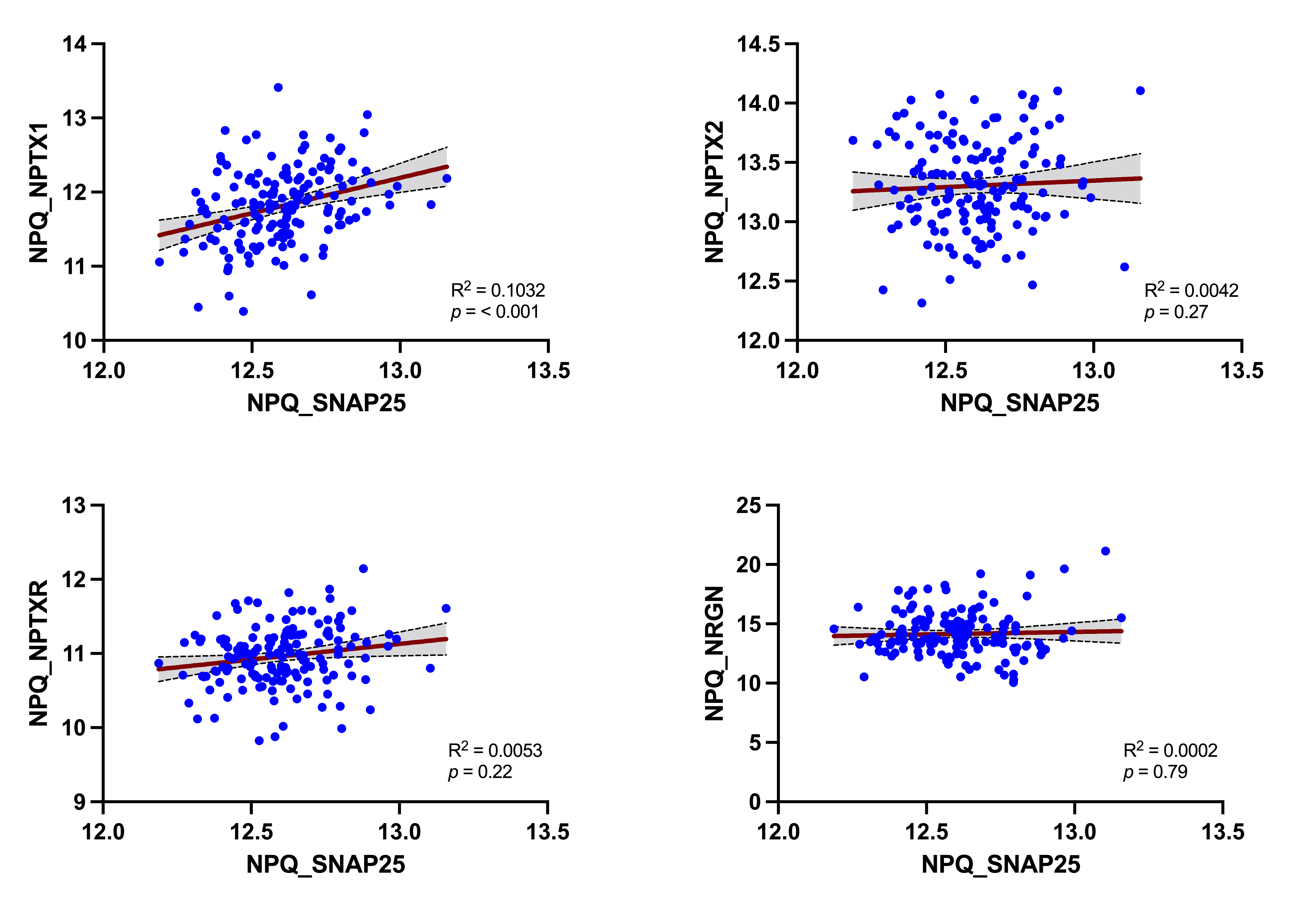

**Supplementary Fig. 4s.** Correlations between core plasma synaptic markers. In AD patients, SNAP-25 showed a significant positive correlation only with NPTX1 (R² = 0.1032; p = <0.001). Red lines indicate linear regression fits with 95% confidence intervals (grey shading).

*Abbreviations: NRGN Neurogranin, NPTX1* neuronal pentraxin 1, *NPTX2* neuronal pentraxin 2, *NPTXR* neuronal pentraxin receptor*, SNAP-25* synaptosomal-associated protein 25; *NPQ, NULISA Protein Quantification*

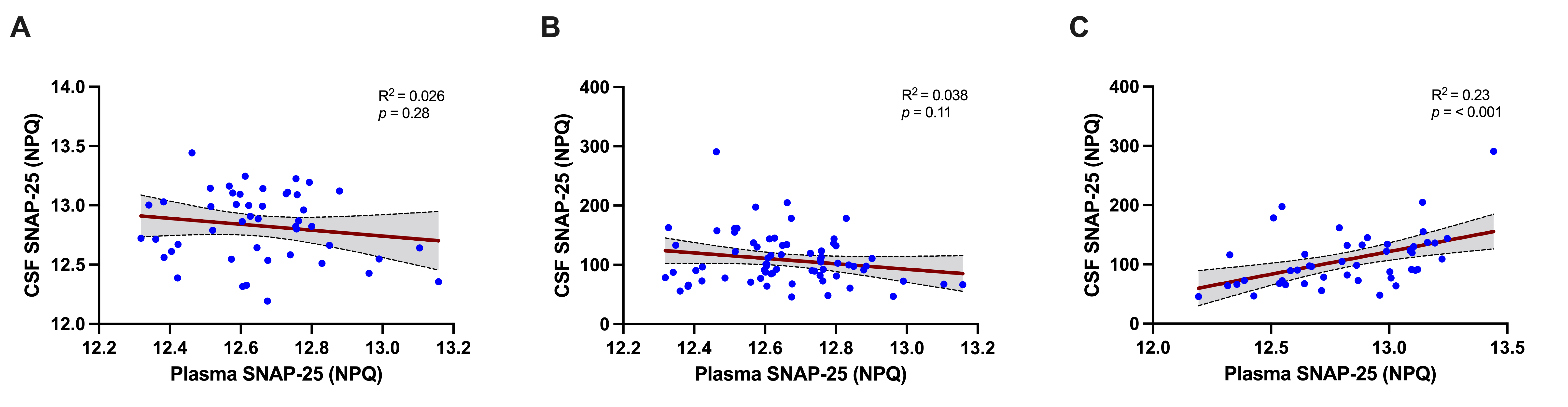

**Supplementary Fig. 5s.** Cross-platform comparison of SNAP-25 measurements. (A) Plasma vs CSF SNAP-25 measured with NULISA (B) Plasma SNAP-25 (NULISA) vs CSF SNAP-25 (Simoa) (C) CSF SNAP-25 measured with Simoa vs NULISA. In all panels, individual data points are displayed in blue, with regression line and 95% confidence interval shown in black and grey, respectively.

*Abbreviations: Simoa Single molecule array, NULISA NUcleic acid Linked Immuno-Sandwich Assay, SNAP-25 synaptosomal-associated protein 25, NPQ, NULISA Protein Quantification.*
